# Intraoperative classification of glioblastoma through near real-time stimulated Raman scattering microscopy

**DOI:** 10.64898/2025.11.28.25341141

**Authors:** Jakob Straehle, Nicolas Neidert, Giulia Villa, Pierre Scheffler, Zeynep Mercan, David Reinecke, Anna-Katharina Meissner, Abdulkader Al Shughri, Roland Goldbrunner, Georg Widhalm, Lisa I. Körner, Barbara Kiesel, Jessica Makolli, Thomas Roetzer-Pejrimovsky, Karl Rössler, Lasse Dührsen, Ulrich Schüller, Richard Drexler, Philipp Karschnia, Adelheid Wöhrer, Christian Freyschlag, Volker A. Coenen, Daniel Erny, Roman Sankowski, Franz Ricklefs, Roland Roelz, Daniel Delev, Felix Sahm, Oliver Schnell, Marco Prinz, Christian W. Freudiger, Volker Neuschmelting, PRECISE Consortium, Jürgen Beck, Dieter Henrik Heiland

## Abstract

Glioblastoma is a highly malignant brain tumor in which maximal safe resection is associated with improved survival, yet the oncological benefit of resection varies by molecular subtype. Recent work has shown that DNA methylation-defined subtypes, particularly receptor tyrosine kinase (RTK) I and II, benefit from complete CE (contrast-enriched) resection compared to mesenchymal tumors, highlighting the need for pre- or intraoperative tools that guide resection based on tumor biology. Here, we present ***iSTAMP*** (**i**ntraoperative **S**patially-informed **T**umor **A**rchitecture **M**apping and **P**rofiling) a real-time, label-free molecular classification framework using stimulated Raman scattering microscopy and graph-based deep learning to predict glioblastoma epigenetic subtypes intraoperatively (within 5-7 minutes). Across 1,295 intraoperative tissue samples from 236 patients profiled with EPIC methylation arrays, our graph attention network achieved high predictive performance for all major subtypes (AUC range 0.88-0.99), with spatially stable predictions across tumor regions. RTK subtypes, but not mesenchymal tumors, showed significant survival benefit from GTR (HR = 0.42, p = 6.1 ×10^-6^). Explainable AI methods revealed subtype-specific histopathological features, including necrosis and macrophage infiltration in mesenchymal tumors versus glio-fibrillary matrix or axon-rich regions in RTK tumors. Spatial transcriptomic validation confirmed cellular correlates with defined subtype specific SRH features. These findings support the integration of Raman-based molecular diagnostics into intraoperative workflows to guide biologically informed surgical strategies in glioblastoma.

## Introduction

Glioblastoma (GB) is the most aggressive primary brain tumor in adults, marked by highly infiltrative growth, intrinsic molecular heterogeneity, and inevitable recurrence. Reported by the RANO resect group, maximal safe surgical resection as defined on MR imaging remains the cornerstone of initial therapy and is strongly associated with improved survival ^1,2^. However, achieving extensive resection, particularly supramarginal resection into the infiltrative tumor margins, is associated with increased risk of potential neurological morbidity. Balancing oncological efficacy with functional preservation remains a central challenge in glioblastoma surgery. In recent years, epigenetic classification has reshaped our understanding of glioblastoma biology, stratifying tumors into distinct molecular subtypes with differing biology and clinical behavior^3^. The Heidelberg DNA methylation classifier defines three primary subgroups in IDH-wildtype glioblastoma: receptor tyrosine kinase 1 (RTK 1), RTK 2, and mesenchymal-like (Mes)^4^. Although these subtypes do not differ significantly in overall survival when considered in isolation, emerging evidence suggests subtype-specific benefits from surgical intervention. Patients with RTK subtypes show a survival benefit from gross total resection (GTR), compared to patients with mesenchymal tumors, often characterized by a more immune-enriched and infiltrative phenotype, that demonstrate less benefit from aggressive surgical approaches^5^. Additionally, recent studies have introduced the concept of “neural-high” versus “neural-low” methylation contexts, linking peritumoral neural integration with poor survival and higher need for complete resection, further emphasizing the clinical relevance of intraoperative molecular diagnostics^6^. The ability to identify glioblastoma subtypes during surgery could guide judicious surgical decision-making and offer real-time, personalized decisions regarding the extent of resection. However, current intraoperative diagnostic methods are limited. While real-time nanopore sequencing offers accurate DNA methylation profiling, it is constrained by lengthy workflows, cost, and infrastructure requirements, limiting its routine intraoperative application ^7,8^. Stimulated Raman scattering, a rapid, label-free imaging modality, has emerged as a promising alternative. Capable of producing high-resolution histology-like images within minutes^9^, Raman-based imaging has already been applied to classify IDH mutation status, 1p/19q codeletion, ATRX deletion ^10,11^, and tumor infiltration in situ ^12–19^. Nevertheless, traditional deep learning approaches for tumor subtyping, such as convolutional neural networks and vision transformers, often focus on isolated image patches or small regions and fail to capture the broader architectural context of the tumor microenvironment. To overcome these limitations, we employed *iSTAMP*, a spatially informed framework using graph-based deep learning from tumor environment-associated graphs which model local neighborhoods within the tissue architecture and incorporate both node-level image embeddings and edge-based spatial relationships. This approach allows for biologically interpretable predictions through integrated gradients and attention-based attribution, linking model decisions to morphologically meaningful structures and demonstrated high accuracy in transfer learning in our recent work ^20^.

In this study, we present a multicenter analysis of intraoperative Raman histology in 236 patients with glioblastoma profiled using DNA methylation arrays (EPIC, Illumina). We combined self-supervised learning with a graph attention neural network (GAT) trained on 906 intraoperative biopsies to predict epigenetic subtypes in real time. We demonstrate that subtype predictions are accurate, spatially stable across tumor regions, and explainable at the level of histological and cellular features. Finally, we integrate our predictions with spatial transcriptomics to link Raman-detectable features with underlying tumor cell phenotypes and microenvironmental signatures. This work provides a foundation for real-time, interpretable molecular diagnostics in neurosurgical oncology and supports the implementation of biologically guided, personalized resection strategies.

## Results

### Epigenetic RTK-subtypes are associated with survival benefit from gross total resection

Building on recent findings that intraoperative molecular profiling may inform the extent of resection in glioblastoma, we sought to assess whether the survival benefit associated with greater extent of resection varies across epigenetically defined subgroups. Specifically, we hypothesized that receptor tyrosine kinase (RTK) subtypes 1 and 2 derive disproportionate benefit from gross total resection (GTR), whereas mesenchymal tumors may be less responsive to cytoreductive strategies as demonstrated recently ^6^. To this end, we analyzed a multi-institutional cohort of 283 patients (236 patients plus 46 patients without SRH imaging) with IDH-wildtype glioblastoma profiled using methylation arrays and stratified by the RANO resect classification^2^. We first evaluated whether epigenetic subgroup alone impacts prognosis. In Kaplan-Meier and Cox regression analyses (Figure 1a), patients with RTK1 tumors exhibited a significantly higher risk of death compared to those with mesenchymal tumors (HR = 1.49, 95% CI: 1.04–2.14, P = 0.030), whereas the RTK 2 group showed no significant difference (HR = 1.17, 95% CI: 0.83–1.64, P = 0.37). The overall prognostic performance of the subgroup model was modest (concordance index = 0.542). We next examined the prognostic impact of surgical resection independently of molecular subtype (Figure 1b). Both GTR and partial tumor resection (PTR) were associated with markedly improved survival relative to subtotal resections (RANO IIIa/IIIb). GTR reduced the hazard of death by 97.8% (HR = 0.022, 95% CI: 0.006–0.080, P = 6.2 × 10^−9^), and PTR by 95.0% (HR = 0.050, 95% CI: 0.014–0.177, P = 3.3 × 10^−6^). The model achieved a concordance of 0.633 and was highly significant overall (likelihood ratio test = 41.5, P = 1 × 10^−9^), confirming the well-established survival benefit of maximal resection. To understand whether these effects are independent, we performed multivariable Cox regression incorporating epigenetic subgroup, extent of resection, and MGMT promoter methylation status (Figure 1c). RTK 1 tumors remained independently associated with shorter survival (HR = 1.49, 95% CI: 1.02–2.18, P = 0.039), while RTK 2 tumors did not differ from the mesenchymal reference (HR = 1.13, 95% CI: 0.79–1.61, P = 0.50). Both GTR (HR = 0.028, 95% CI: 0.008–0.10, P = 4.1 × 10^−8^) and PTR (HR = 0.058, 95% CI: 0.016–0.20, P = 9.4 × 10^−6^) remained strong independent predictors of improved outcome, while unmethylated MGMT promoter status was associated with poorer survival (HR = 1.37, 95% CI: 1.02–1.84, P = 0.038). The full model demonstrated robust prognostic accuracy (concordance index = 0.659, likelihood ratio test = 48.2, P = 3 × 10^−9^). We then tested our core hypothesis that the benefit of GTR is specific to epigenetic context. Different recent epigenetic classification demonstrated clinical relevance. For the majority of classes (MGMT, Heidelberg Class and Neural Class) no clear association was demonstrated in our cohort, Figure 1d. Following the major diagnostic classifier (Heidelberg Class), patients were dichotomized into RTK (combining RTK 1 and 2) and Mesenchymal groups, and subgroup-specific Cox regressions were performed (Figure 1e– f). In mesenchymal tumors (Figure 1e), GTR conferred a non-significant survival advantage (HR = 0.67, 95% CI: 0.38–1.16, P = 0.15) with limited model concordance (C-index = 0.581). In contrast, RTK tumors (Figure 1f) demonstrated a pronounced and statistically robust benefit from GTR (HR = 0.42, 95% CI:

**Figure 1.**
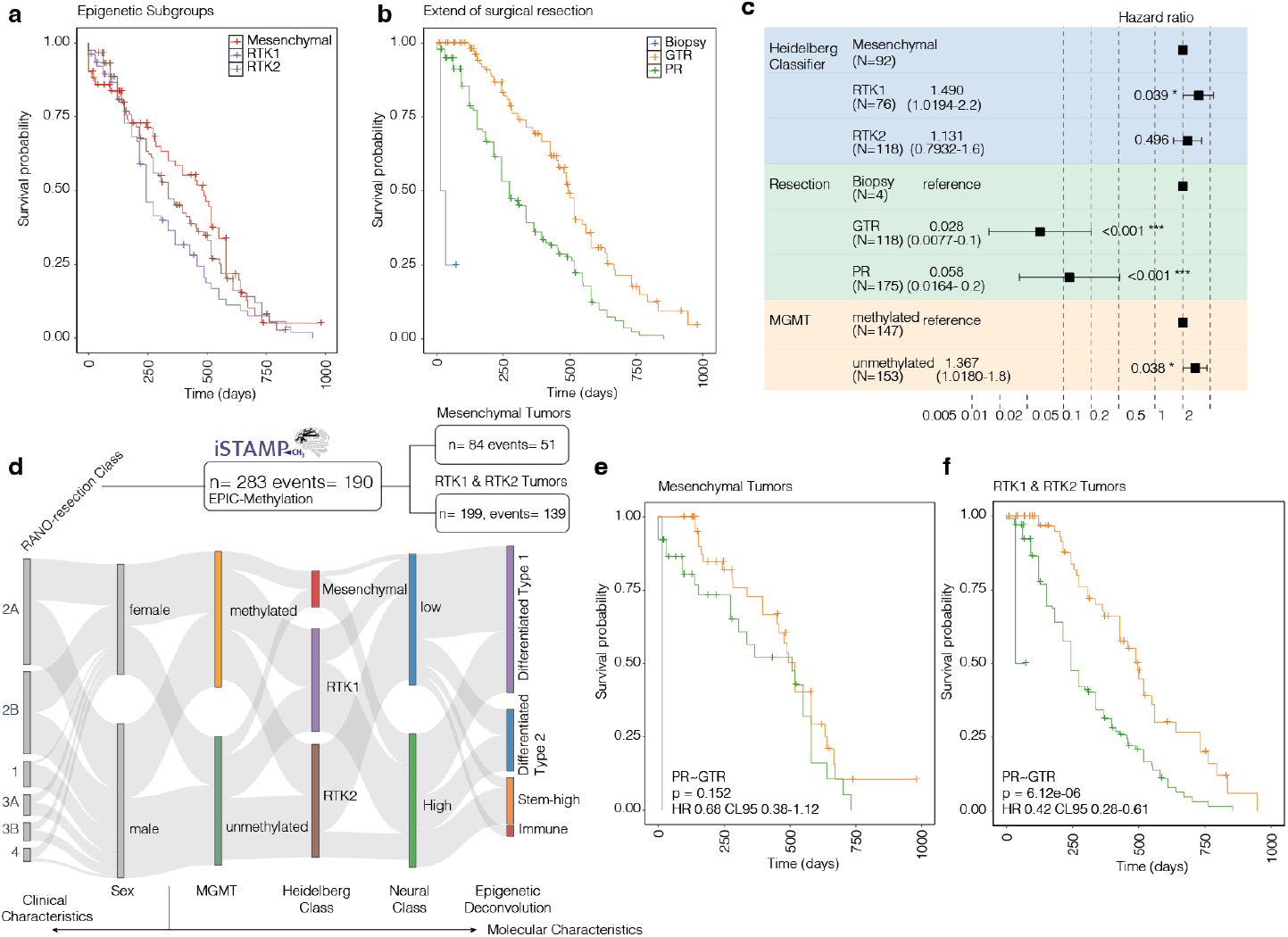
a) Kaplan-Meier survival plot of the epigenetic subgroups RTK 1 (purple) RTK 2 (brown) and mesenchymal (red). b) Kaplan-Meier survival plot of the extend of resection strategies with only biopsy in blue, partial resection (PR) defined as RANO resect group IIIa/b, and gross-total resection including RANO resect group I and IIa/b. c) Forest plot of the cox-regression analysis demonstrating that RTK tumors (RTK1) and unmethylated MGMT promoter show worse outcome. d) Overview of the distribution of clinical and epigenetic classification systems. e) Kaplan-Meier survival plot of the epigenetic subgroup mesenchymal stratified by the extent of resection groups. f) Kaplan-Meier survival plot of the epigenetic subgroup RTK1 and RTK2 stratified by the extent of resection groups.

0.28–0.61, P = 6.1 × 10^−6^) with stronger model performance (C-index = 0.622). These findings support the concept of molecularly guided neurosurgery, in which the aggressiveness of resection may be tailored based on tumor-intrinsic biology. Altogether, our data suggest that maximal surgical resection in glioblastoma offers differential benefit depending on the epigenetic profile of the tumor. While resection improves survival in most patients, the magnitude of benefit appears restricted to RTK tumors, and may be limited in mesenchymal subtypes. These findings underscore the translational potential of intraoperative DNA methylation profiling to optimize neurosurgical strategies on a patient-specific basis.

### Intraoperative profiling of epigenetic glioblastoma subgroups by Raman Spectroscopy

Recent advances in intraoperative molecular diagnostics, including real-time nanopore sequencing and Raman spectroscopy, have opened new avenues for identifying biologically distinct tumor subtypes during neurosurgical procedures. While nanopore sequencing enables accurate, real-time annotation of DNA fragments and epigenetic subgroups, its use in routine clinical workflows is limited by high costs, labor intensity, and prolonged turnaround times (∼1 h or more)^7,8,21^. As an alternative, label-free Raman-based histology has demonstrated the capacity to distinguish glioma molecular subtypes mutationand tumor cell density rapidly (<5 min) and at low cost ^11^. To explore the potential of Raman spectroscopy for intraoperative epigenetic classification, we established ***PRECISE*** *(Prompt Raman SpEctroscopy*

*Controlled OncologIcal Surgery across Europe)*, a European consortium, comprising ten clinical and research sites across three countries. Across this network, we collected intraoperative tissue samples from 236 patients, yielding a total of 1,295 biopsies that were subjected to stimulated Raman histology (INVENIO Imaging) and DNA methylation profiling using EPICv1 (Illumina) arrays (Figure 2a-b).

**Figure 2.**
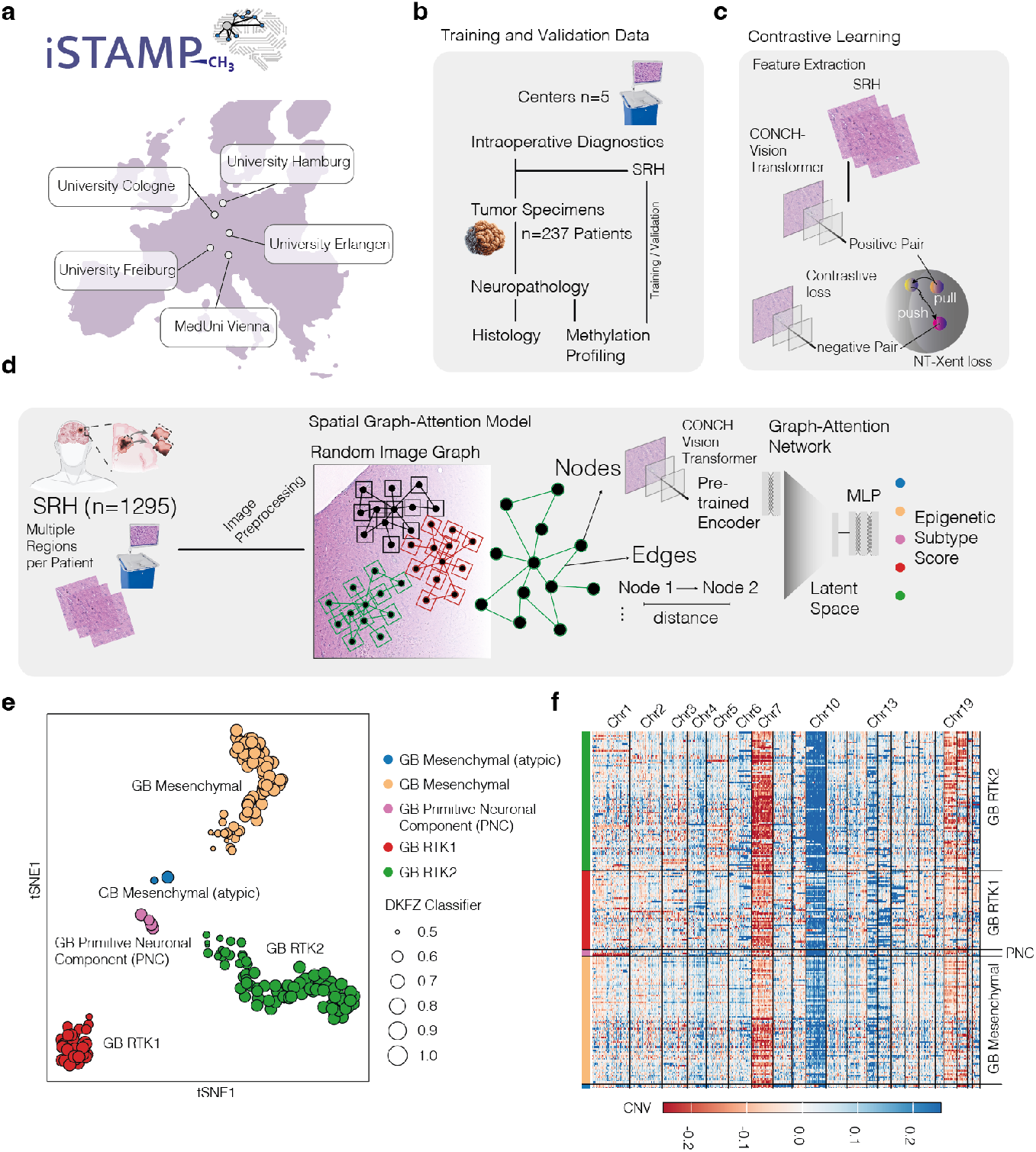
a-d) Overview of the cohort and deep-learning strategy for epigenetic subgroup prediction. e) T-SNE scatter plot of the DKFZ classification scores (v12.8). Size of each dot demonstrate the top classification score. Colors indicate the different subgroups. f) Copy number alteration analysis of the tumor samples (n=236) show common gain in Chr 7 and loss in Chr 10.

To extract molecularly relevant information from stimulated Raman histology (SRH) images, we applied a state-of-the-art deep learning framework adapted from histopathology analysis. Specifically, we utilized CONCH^22^, a pretrained histological foundation model, which we fine-tuned in an unsupervised manner on our SRH dataset to generate low-dimensional embeddings for individual tissue patches (Figure 2c). These embeddings served as the basis for a graph-based representation of each image. Building on prior findings from our group that highlighted the importance of spatial tissue context for molecular inference from histology and transcriptomics^20^, we modeled local tissue architecture by constructing random graph neighborhoods across each SRH image. In this graph, each node represented a histological patch defined by its learned embedding, and edges encoded the physical distance between neighboring patches (size 256×256 pixels) (Figure 2d). We trained a graph attention neural network (GAT) to predict the epigenetic subclass score of each sample, leveraging both node-level image features and inter-patch spatial relationships. The final study cohort comprised 236 patients with IDH-wildtype glioblastoma who underwent intraoperative tissue sampling across four academic centers: University Hospital Freiburg (UKF, n = 90), University Hospital Cologne (UKK, n = 48), Medical University of Vienna (MUV, n = 54), and University Hospital Hamburg (UKH, n = 45). DNA methylation profiling assigned tumors to the following molecular subgroups: GB RTK2 (n = 96), GB MES-TYP (n = 86), GB RTK1 (n = 47), GB PNC (n = 5), and GB MES-ATYP (n = 3). *MGMT* promoter methylation was detected in 116 patients (49%), whereas 121 (51%) were unmethylated. The cohort included 103 female (43%) and 134 male (57%) patients, representing a typical glioblastoma cohort. In total, 1,295 scattered Raman histology images were acquired, with each patient contributing between 1 and 12 biopsies. To train and evaluate our classification model, we randomly divided the dataset into a training and validation cohort using a 70:30 split, stratified to balance key covariates including *MGMT* promoter methylation status, sex, and the number of tissue images per clinical center. The resulting training set consisted of 166 patients with 906 images, while the validation set included 71 patients with 389 images. This approach ensured both molecular and demographic representativeness across subsets, allowing robust and generalizable model performance.

### Intraoperative profiling of epigenetic glioblastoma subgroups by Raman Spectroscopy

To evaluate the ability of our graph attention model to classify glioblastoma epigenetic subtypes from intraoperative stimulated Raman histology, we first assessed its performance on the held-out validation cohort. Receiver operating characteristic (ROC) curve analysis demonstrated robust multiclass classification performance, with high area under the curve (AUC) values for all major subtypes (Figure 3a). The model achieved an AUC of 0.905 for GB MES-TYP, 0.993 for GB MES-ATYP, 0.960 for GB PNC, 0.877 for GB RTK1, and 0.892 for GB RTK2, indicating strong discriminatory power across morphologically and molecularly distinct groups. Investigating single subgraphs containing 20-40 nodes (patches 256×256 pixels) and their distance relationships, we found strong classification performance for GB RTK2 and GB RTK1, with misclassifications largely concentrated between adjacent or biologically similar subtypes. Notably, misclassification occurred most frequently in the GB MES-TYP group, which exhibited some overlap with RTK subtypes, likely reflecting morphological diversity in this subgroup (Figure 3b). We next visualized the learned latent representations using t-SNE dimensionality reduction which formed distinct clusters corresponding to ground truth epigenetic subtypes, with tight spatial separation for RTK1, RTK2, and PNC tumors, Figure 3c.

**Figure 3.**
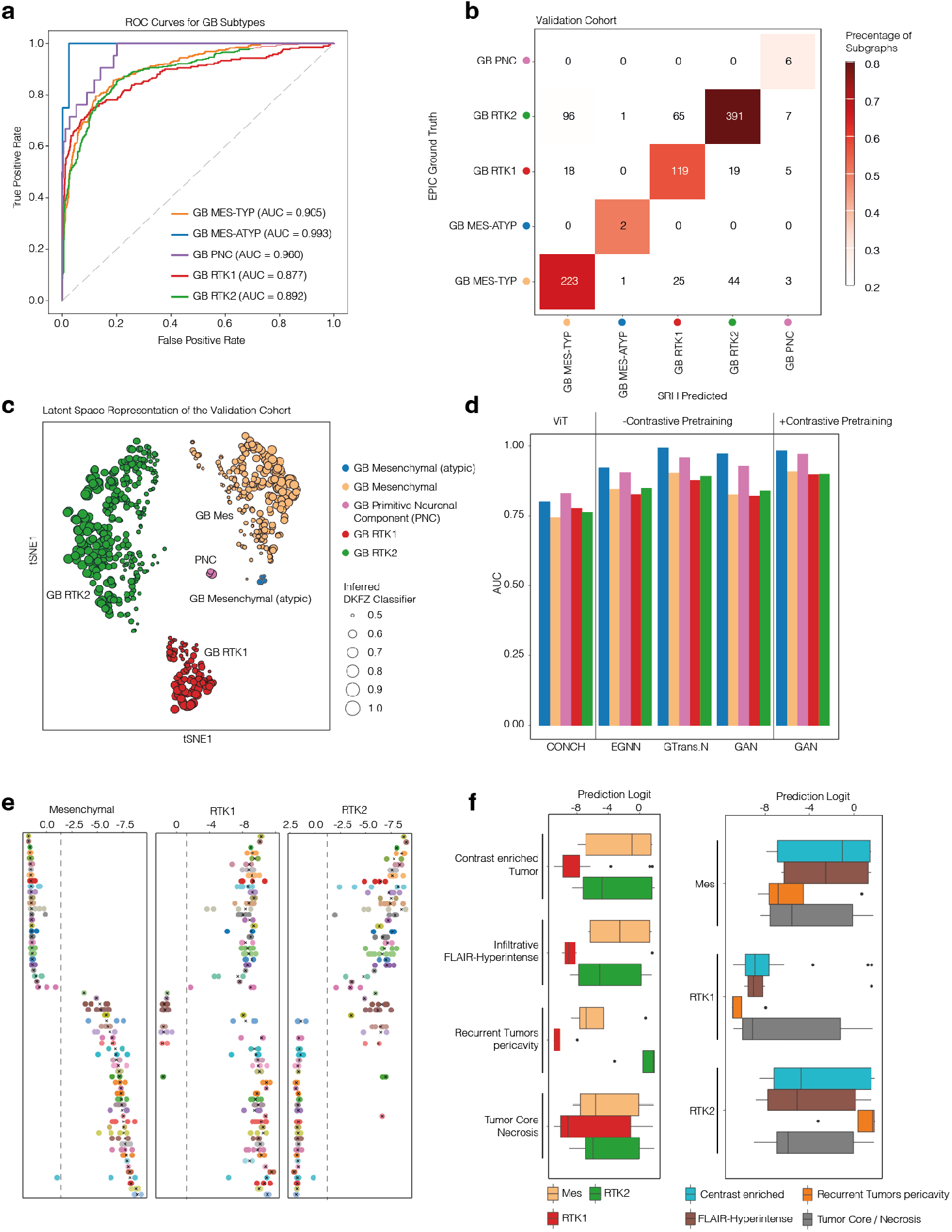
a) Receiver operating characteristic (ROC) curves for subtype classification performance on the validation cohort. The model demonstrated high area under the curve (AUC) values for all major epigenetic glioblastoma subtypes b) Normalized confusion matrix displaying classification accuracy and subtype-specific misclassifications in the validation cohort at single subgraph level. Most errors occurred between biologically related groups, particularly GBM_MES_TYP and GBM_RTK2. c) t-distributed stochastic neighbor embedding (t-SNE) visualization of the latent space learned from Raman histology in the validation cohort. Each dot represents a histological patch, colored by true epigenetic subtype, and scaled by classifier confidence (Heidelberg Classifier score). Distinct subtype clusters reflect strong representation learning. d) Comparison of model performance (AUC) across different architectures with and without contrastive pretraining. Models included Contrastive Learning of Neighborhoods for Classification in Histology (CONCH), Equivariant Graph Neural Network (EGNN), Graph Transformer Network (GTrans.N), and Graph Attention Network (GAT). Contrastive pretraining improved performance across all models, with the GAN-based architecture achieving the highest accuracy. e) Distribution of prediction logits for Mesenchymal, RTK1, and RTK2 glioblastoma subtypes across multiple spatially resolved biopsies per patient. Each dot represents a biopsy sample, color-coded by individual patient. Prediction scores show limited intra-patient variability, with the majority of samples falling within a narrow confidence interval, indicating spatially stable classification. The cross indicates the median logit. f) Prediction logit distributions stratified by MRI-defined tumor regions: contrast-enhancing tumor, infiltrative FLAIR-hyperintense zones, recurrent tumors adjacent to the resection cavity, and necrotic tumor core. Left: Logit values grouped by epigenetic subtype. Right: Logit values grouped by anatomical tumor region, Right: Logit values grouped by epigenetic subtype. Mesenchymal and RTK2 tumors show consistent prediction strength across all anatomical compartments, while RTK1 tumors display generally lower scores, likely due to lower representation in the training dataset. Boxplots represent interquartile range (IQR), with whiskers indicating 1.5× IQR and individual dots representing outliers.

The classifier confidence, as inferred by Heidelberg Classifier scores, correlated with intra-cluster compactness, supporting the reliability of spatial embedding for subtype prediction. To assess the benefit of model architecture and pretraining strategies, we compared several deep learning frameworks under different training conditions. These included Contrastive Learning of Neighborhoods for Classification in Histology (CONCH), Equivariant Graph Neural Networks (EGNN), Graph Transformer Networks (GTrans.N), and Graph Attention Network (GAT). Across all architectures, the integration of contrastive pretraining improved classification performance. The highest overall accuracy across epigenetic subtypes was achieved by our graph attention model trained with contrastive pretraining, underscoring the value of combining spatial context with representation learning.

### Low intra-tumoral heterogeneity of epigenetic predictions

To investigate intra-tumoral heterogeneity in epigenetic subgroup prediction, we analyzed multiple spatially resolved biopsies per patient, annotated according to intraoperative neuronavigation and matched MRI sequences. Tissue regions were classified into contrast-enhancing tumor, infiltrative FLAIR-hyperintense margins, tumor core/necrosis, and recurrent tumor zones adjacent to the resection cavity. We quantified the variance in model-derived prediction logits across biopsies from individual patients and subtypes (Figure 3e). In the majority of cases (92%), intra-patient prediction scores varied within a narrow range typically between 2% and 10% of the 95% confidence interval (CI) of the subtype logit distribution indicating high intra-tumoral consistency in predicted class. Only a single patient demonstrated prediction variability across biopsies that altered the final classification. To assess whether model confidence was influenced by anatomical tumor subregions, we analyzed prediction logits across MRI-defined compartments for each epigenetic subtype (Figure 3f). RTK2 and mesenchymal tumors displayed consistent prediction strengths across all spatial regions, including the necrotic core, infiltrative margins, and contrast-enhancing zones, suggesting robust spatial generalizability of the model. In contrast, RTK1 tumors exhibited lower overall logit scores, likely reflecting underrepresentation of this subgroup in the training cohort; however, no significant intra-subgroup differences were observed across anatomical regions. These results indicate that Raman-based subtype predictions are spatially stable across heterogeneous tumor zones and support the potential for single-biopsy diagnosis using intraoperative imaging guidance.

### Explainable AI-tools to investigate spatial architecture of epigenetic subgroups

To further investigate the histological correlates underpinning Raman-based epigenetic classification, we applied explainable AI methods to interpret the model’s spatial decision-making. Specifically, we used attention scores from the final attention head of the graph attention network to identify highly weighted nodes and integrated gradients to quantify the contribution of individual patches to subtype prediction. Subgraphs were extracted from spatially localized regions of the SRH images, with each node representing a histological patch and each edge encoding its spatial relationship to neighboring patches. Top predictive subgraphs for each epigenetic class were visualized and analyzed morphologically (Figure 4a-c). In mesenchymal tumors, predictive subgraphs were enriched for necrotic regions and hypervascularized areas, consistent with known histopathological features of this subtype (Figure 4a). RTK1-associated subgraphs displayed prominent (glia-) fibrillar structures and regions populated by reactive astrocytes, suggesting a gliotic microenvironment (Figure 4b). RTK2-associated subgraphs were dominated by tightly packed, high-density cellular areas with saturated (lipid) fiber structures (axons), reflective of the hyperproliferative nature of this subgroup (Figure 4c). These spatially resolved attention maps and feature attributions confirm that the model learns biologically meaningful morphological patterns consistent with human expert interpretation, thereby reinforcing the interpretability and robustness of Raman-based deep learning classification of glioblastoma subtypes.

**Figure 4.**
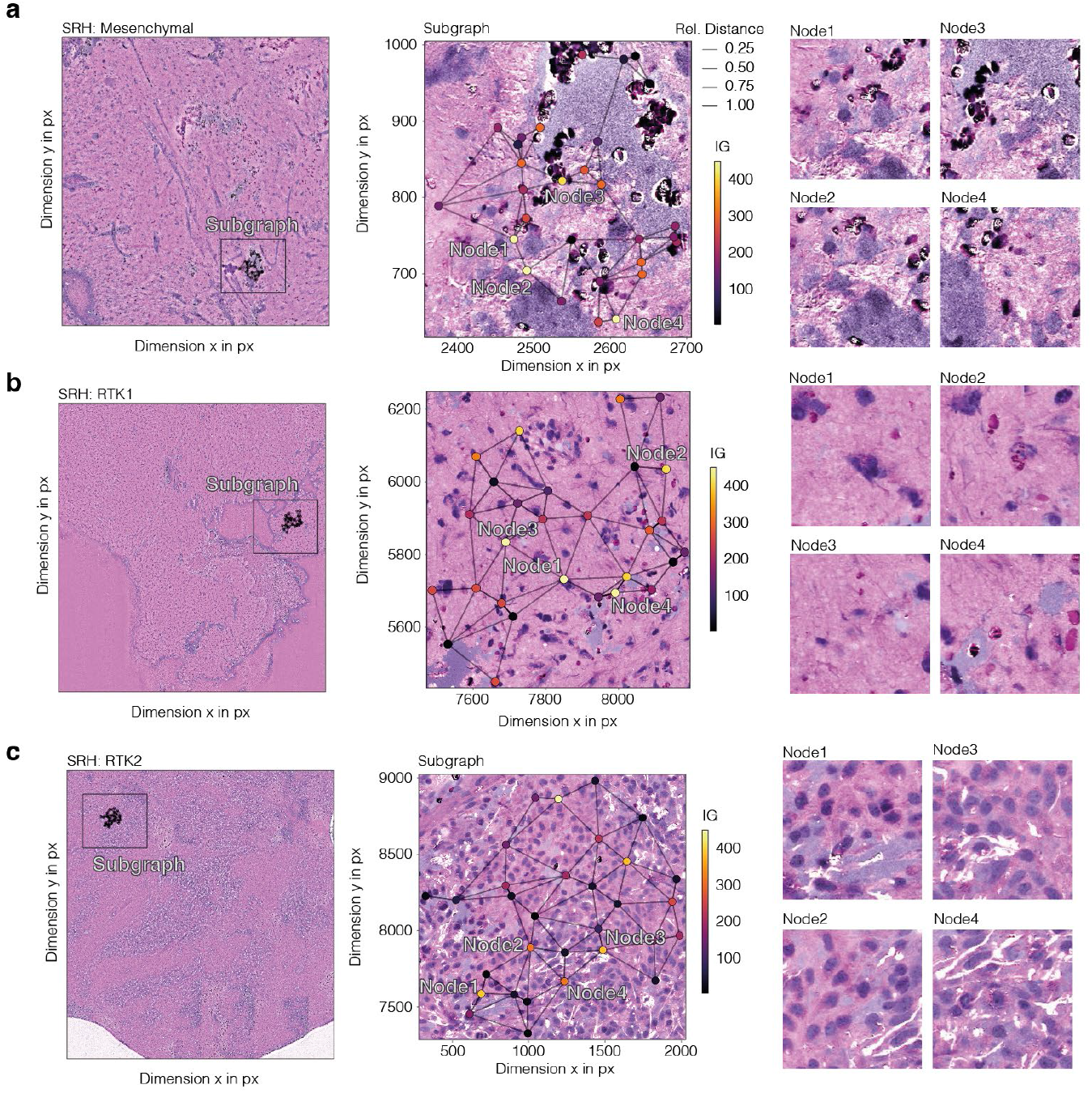
a–c, Representative stimulated Raman histology images and corresponding subgraph visualizations from mesenchymal (a), RTK1 (b), and RTK2 (c) glioblastomas. Left: Full tissue section with location of the analyzed subgraph highlighted. Middle: Enlarged view of the subgraph with attention-weighted graph structure. Nodes represent individual histological patches, and edges encode relative spatial distance (line width). Color indicates feature importance calculated by integrated gradients (IG). Right: Zoomed views of individual high-importance nodes within the subgraph (Node1–Node4), revealing morphologies associated with subtype classification. Mesenchymal subgraphs exhibit necrosis and hypervascularization, RTK1 shows reactive gliosis and gliofibrillar regions, and RTK2 is characterized by high cellular density and saturated fiber (axonal) networks.

### Integration of Scattered Raman Spectroscopy and spatially resolved transcriptomics

To understand the transcriptional and cellular context underlying Raman-based epigenetic subtype predictions, we established an integrative spatial profiling pipeline combining SRH with spatially resolved transcriptomics.

Freshly excised biopsies were first imaged with SRH and then embedded in OCT for array-based transcriptomic profiling using the 10x Genomics Visium platform at 55 μm resolution (n = 9 patients). The tissue sections were cryosectioned, stained, and spatially co-registered with the original SRH images using aligned H&E scans (Figure 5a). SRH images were then processed to extract spatial graphs and predict epigenetic subtypes using the trained model. To assess the biological correlates of prediction strength, we computed prediction logits across the tissue section and compared them to cell-type composition inferred from spatial transcriptomics (Figure 5b-d). We found that regions with high prediction confidence for the mesenchymal subtype were consistently enriched for myeloid-lineage cells, in particular tumor-associated macrophages (p_adj_=2.34 x 10^-16^) (TAMs) (Figure 5e). These findings align with prior studies linking mesenchymal glioblastomas to a highly inflamed, immune-infiltrated tumor microenvironment. In contrast, regions predicted as RTK1 showed enrichment of astrocyte-like tumor cells (p_adj_=1.762 x 10^-7^), which histologically corresponded to darker, protein-rich fibrillar structures observed in SRH images. This pattern likely reflects a denser tumor-tumor communication network, previously associated with astrocytic-like tumor architecture. Regions with high RTK2 prediction scores were dominated by oligodendrocyte precursor cell (OPC)-like tumor phenotypes (p_adj_=4.24 x 10^-4^) and a greater abundance of normal glial populations, including OPCs, astrocytes, and mature oligodendrocytes. The presence of lipid-rich fiber structures in RTK2 regions likely reflects myelinated axons or residual myelin debris, providing a structural/histological basis for the identified SRH-associated features by the model. Altogether, this multimodal integration links chemical imaging with high-resolution gene expression and cellular composition, offering biological interpretability to deep learning–based prediction and revealing subtype-specific structural and cellular signatures within the glioblastoma microenvironment.

**Figure 5.**
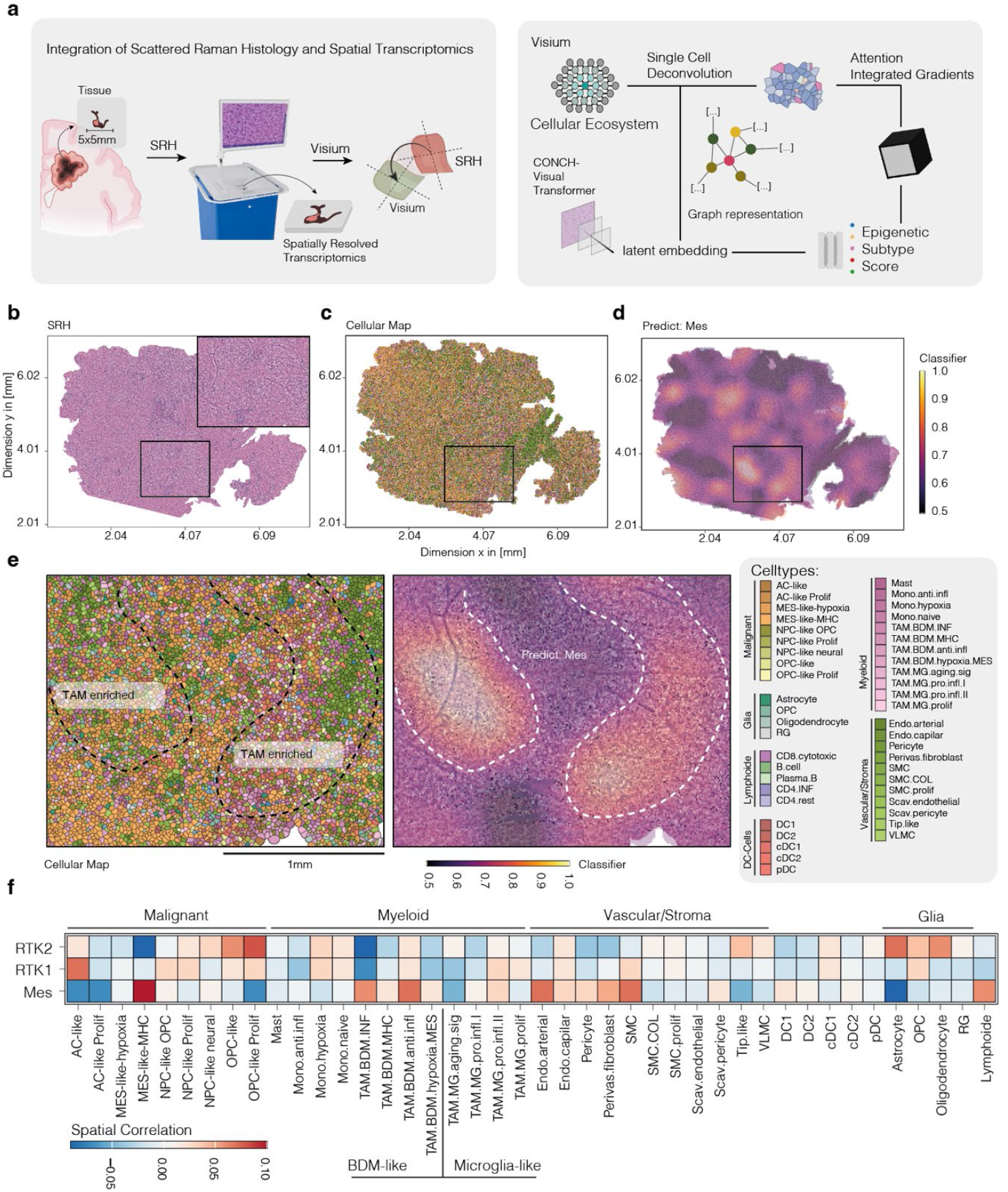
a) Schematic of the multimodal workflow. Fresh glioblastoma tissue was imaged using SRH, embedded in OCT, cryosectioned, and processed using the Visium spatial transcriptomics platform. SRH images were co-registered to Visium-derived H&E scans. Preprocessed SRH graphs were used for subtype prediction and explainability analyses via attention and integrated gradients. B) SRH image of a representative tumor section with regions of interest marked. C) Corresponding cell-type composition map derived from spatial transcriptomic deconvolution, showing the spatial distribution of inferred cell types. d) Spatial heatmap of mesenchymal prediction logits across the same section. e) Magnified view of TAM-enriched areas corresponding to high mesenchymal prediction scores, aligned with the SRH image. F) Heatmap summarizing cell-type composition across tumor regions predicted as RTK1, RTK2, or mesenchymal, showing differential enrichment of myeloid, glial, stromal, and malignant subtypes. RTK2 regions show OPC-like and oligodendrocyte enrichment; RTK1 is associated with astrocytic-like tumor cells; and mesenchymal regions are characterized by TAM and stroma enrichment.

## Discussion

In this study, we demonstrate that Raman-based intraoperative imaging, combined with spatially informed deep learning, enables accurate classification of glioblastoma subtypes as defined by DNA methylation profiling. Using a European multi-institutional cohort and over 1,200 intraoperative biopsies, we confirm that receptor tyrosine kinase (RTK) subtypes, particularly RTK1 and RTK2, derive a significant survival benefit from gross total resection, whereas mesenchymal tumors exhibit limited resection-related survival improvement. This observation underscores the clinical importance of molecularly guided neurosurgical strategies. While nanopore sequencing has previously been proposed for intraoperative methylation-based diagnosis, its time, cost, and infrastructure requirements limit its broader clinical applicability^7,87,8.^ In contrast, stimulated Raman histology combined with deep learning provides a label-free, low-cost alternative capable of rapid inference within surgical timeframes ^12–19^. In our cohort, we reached a prediction end-to-end of SRH images in approximately 4.5 minutes. By generating high-dimensional image embeddings and modeling local spatial architecture through graph attention networks, our approach achieves robust subtype prediction accuracy and captures biologically meaningful histomorphological features. Importantly, our model demonstrated low intra-tumoral prediction heterogeneity, with consistent subtype calls across diverse tumor regions including infiltrative margins, necrotic zones, and contrast-enhancing cores. This finding supports the reliability of single-biopsy prediction during surgery. Using explainable AI tools such as integrated gradients and attention maps, we identified subtype-specific histopathological features that aligned with known biological hallmarks such as necrosis and TAM enrichment in mesenchymal tumors, fibrillar gliotic regions in RTK1, and high glial content and myelinated debris in RTK2. Integrating Raman imaging with spatial transcriptomics further revealed that prediction strength correlates with distinct cellular environments. Mesenchymal predictions were linked to myeloid-rich, TAM-dominated regions, while RTK2 predictions aligned with OPC-like tumor cells and non-malignant glia. These spatial correlations provide mechanistic insight into the biochemical signatures captured by Raman spectroscopy and explain the model’s capacity for subtype discrimination. Altogether, our results highlight the potential of real-time chemical imaging coupled with interpretable AI as a viable alternative to sequencing-based approaches for intraoperative glioblastoma subclassification. This strategy holds promise for improving intraoperative decision-making and tailoring the extent of resection based on tumor biology. Future work will focus on prospective validation and real-time clinical deployment, as well as extending this approach to other tumor types and molecular phenotypes.

## Data Availability

All data produced in the present study are available upon reasonable request to the authors

## Acknowledgements

We thank B. Baumer, O. Müller, J. Göldner, V. Sacalean, S. Heinemann and K. Kolbe for excellent technical support. We would like to acknowledge the use of BioRender.com (licenses: “Student Plan” by D.H.H.) for the creation of illustrations present in this publication.

